# Rapid-CNS^2^: Rapid comprehensive adaptive nanopore-sequencing of CNS tumors, a proof of concept study

**DOI:** 10.1101/2021.08.09.21261784

**Authors:** Areeba Patel, Helin Dogan, Alexander Payne, Philipp Sievers, Natalie Schoebe, Daniel Schrimpf, Damian Stichel, Nadine Holmes, Philipp Euskirchen, Jürgen Hench, Stephan Frank, Violaine Rosenstiel-Goidts, Miriam Ratliff, Nima Etminan, Andreas Unterberg, Christoph Dieterich, Christel Herold-Mende, Stefan M Pfister, Wolfgang Wick, Matthias Schlesner, Matthew Loose, Andreas von Deimling, Martin Sill, David TW Jones, Felix Sahm

## Abstract

**Background:** The 2021 WHO classification of central nervous system tumors includes multiple molecular markers and patterns that are recommended for routine diagnostic use in addition to histology. Sequencing infrastructures for complete molecular profiling require considerable investment, while batching samples for sequencing and methylation profiling can delay turnaround time. We introduce RAPID-CNS^2^, a nanopore adaptive sequencing pipeline that enables comprehensive mutational, methylation and copy number profiling of CNS tumours with a single, cost-effective sequencing assay. It can be run for single samples and offers highly flexible target selection that can be personalized per case with no additional library preparation.

**Methods:** Utilizing ReadFish, a toolkit enabling targeted nanopore sequencing without the need for library enrichment, we sequenced DNA from 22 diffuse glioma samples on a MinION device. Target regions comprised our Heidelberg brain tumor NGS panel and pre-selected CpG sites for methylation classification using an adapted random forest classifier. Pathognomonic alterations, copy number profiles, and methylation classes were called using a custom bioinformatics pipeline. The resulting data were compared to their corresponding standard NGS panel sequencing and EPIC methylation array results.

**Results:** Complete concordance with the EPIC array was found for copy number profiles. The vast majority (94%) of pathognomonic mutations were congruent with standard NGS panel-seq data. *MGMT* promoter status was correctly identified in all samples. Methylation families from the random forest classifier were detected with 96% congruence. Among the alterations decisive for rendering a WHO 2021 classification-compatible integrated diagnosis, 97% of the alterations were consistent over the entire cohort (completely congruent in 19/22 cases and sufficient for unequivocal diagnosis in all 22 samples).

**Conclusions:** RAPID-CNS^2^ provides a swift and highly flexible alternative to conventional NGS and array-based methods for SNV/InDel analysis, detection of copy number alterations, target gene methylation analysis (e.g. *MGMT*) and methylation-based classification. The turnaround time of ∼5 days for this proof-of-concept study can be further shortened to < 24h by optimizing target sizes and enabling real-time computational analysis. Expected advances in nanopore sequencing and analysis hardware make the prospect of integrative molecular diagnosis in an intra-operative setting a feasible prospect in future. This low-capital approach would be cost-efficient for low throughput settings or in locations with less sophisticated laboratory infrastructure, and invaluable in cases requiring immediate diagnoses.

---

Molecular markers are now unequivocally a requirement for integrative brain tumor diagnostics. The 2021 WHO classification of CNS tumors substantially increases the set of genes required in routine evaluation, and significantly increases the relevance of DNA methylation analysis in the diagnostic process [10]. Multiple approaches are available for such analyses. However, neuropathology labs cannot rely on current off-the-shelf products, since these do not cover all genes relevant for neuro-oncology, while including large target regions that are dispensable. Thus, custom assays have typically been set-up in neuropathology labs where the equipment for next-generation sequencing (NGS) is available. In turn, the advantages of custom neuropathology NGS panels can only be efficiently exploited when case numbers are sufficient for batchwise processing. Labs with lower specimen submission numbers hence may have to pool samples over multiple weeks. Here we introduce RAPID-CNS^2^ - a custom neurooncology molecular diagnostic workflow using third generation sequencing for parallel copy-number profiling, mutational and methylation analysis that is highly flexible in target selection, runs efficiently on single samples, and can be initiated immediately upon receipt of frozen sections.

Nanopore sequencing has an advantage over current NGS methods in terms of longer read lengths, shorter and easier library preparation protocols, ability to call base modifications natively from extracted nucleic acids, real time analysis, and portablility of sequencing devices – all at relatively low cost [3]. However, smaller devices like the MinION yield low-coverage data when run genome-wide, that makes it difficult to detect pathognomonic genetic alterations or hard-to-map regions like the *TERT* promoter [6]. Nanopore provides a “ReadUntil” adaptive sampling toolkit that can reject reads in real-time during sequencing [7]. ReadFish harnesses this functionality to enable targeted adaptive sequencing with no additional steps in library preparation [9]. This considerably increases coverage over “target” regions by real-time enrichment during sequencing, to allow confident detection of clinically relevant alterations.

RAPID-CNS^2^ leverages adaptive nanopore sequencing through ReadFish and is run here as a proof-of-concept using a portable MinION device. We formulated target regions covering the Heidelberg brain tumour NGS panel and CpG sites required for methylation-based classification [4, 11]. We performed ReadFish-based sequencing on 22 diffuse glioma samples that had previously undergone brain tumor NGS panel and Infinium MethylationEPIC array (EPIC) analysis [2, 4, 11]. Samples were selected to cover a variety of the most clinically-relevant pathognomonic alterations (*IDH1*, 1p/19q codeletion, chr7 gain/chr10 loss, *TERT* promoter, *EGFR* amplification, *CDKN2A/B* deletion, *MGMT* status) and relevant methylation classes identified by conventional methods. Cryoconserved brain tumour tissue was prepared for Nanopore sequencing with the SQK-LSK109 Ligation Sequencing Kit from ONT. Incubation time and other parameters were optimized to improve quality, amount of data generated and on-target rate of the libraries (Supplementary methods). Single samples were loaded onto FLO- MIN106 R9.4.1 flow cells and run on a MinION 1B. ReadFish controlled the sequencing in real-time and was run using a consumer notebook powered by an 8GB NVIDIA RTX 2080 Ti GPU. Samples were sequenced for up to 72 hours. Our selected target regions covered 5.56% of the entire genome. Sequencing time can be reduced to less than 24 hours by further optimizing the size of the targeted regions. Sequenced data was analysed using a bioinformatics pipeline customized for neurooncology targets (which will be available on https://github.com/areebapatel/RAPID-CNS2). SNVs were filtered for clinical relevance by their 1000 genomes population frequency (<0.01) and COSMIC annotations [1, 13, 14]. Copy number alterations were estimated using depth-of-coverage of the mapped reads [12]. Nanopore sequencing provides the additional advantage of natively estimating base modifications from a single DNA sequencing assay. Methylated bases were identified using megalodon, a deep neural network-based modified base caller [8]. Megalodon’s output was used to compute methylation values over targeted CpG sites and assess *MGMT* promoter methylation status. A random forest classifier based on the previously published reference set [4] was trained to predict methylation classes for the samples. *MGMT* promoter methylation status was assigned by averaging methylation values over all CpG sites in the *MGMT* promoter region (Supplementary results). Mean run time from tissue collection to reporting for RAPID-CNS^2^ was < 5 days. Nanopore sequencing considerably reduced library preparation time to 3.5 hours, as opposed to 48 hours for panel sequencing and 72 hours for the EPIC array (Figure 1). Additionally, it merged both data categories (sequencing and methylation) into one lab workflow. Despite the differences in sequencing technology and method-specific data analysis pipelines, congruence of detected SNVs, regardless of clonality and clinical relevance, was 78% (Supplementary data). Importantly, diagnostically relevant, pathognomonic mutations like *IDH1* R132H/S and *TERT* promoter were congruent in 22/22 and 19/22 samples respectively (Figure 1b). In addition, we derived copy-number-plots (CNP) from calculated copy number levels for the Nanopore data (Supplementary figure 1a). Plots generated using Nanopore data displayed markedly better resolution than those obtained using panel sequencing data (Supplementary figure 1b). Complete concordance with EPIC array analysis was found for CNV levels in all samples. RAPID-CNS^2^ also enabled gene-level CNV detection (Supplementary data). Among the alterations decisive for rendering an integrated molecular diagnosis, 217/220 were consistent over the entire cohort (completely congruent in 19/22 cases).

**Figure 1:**
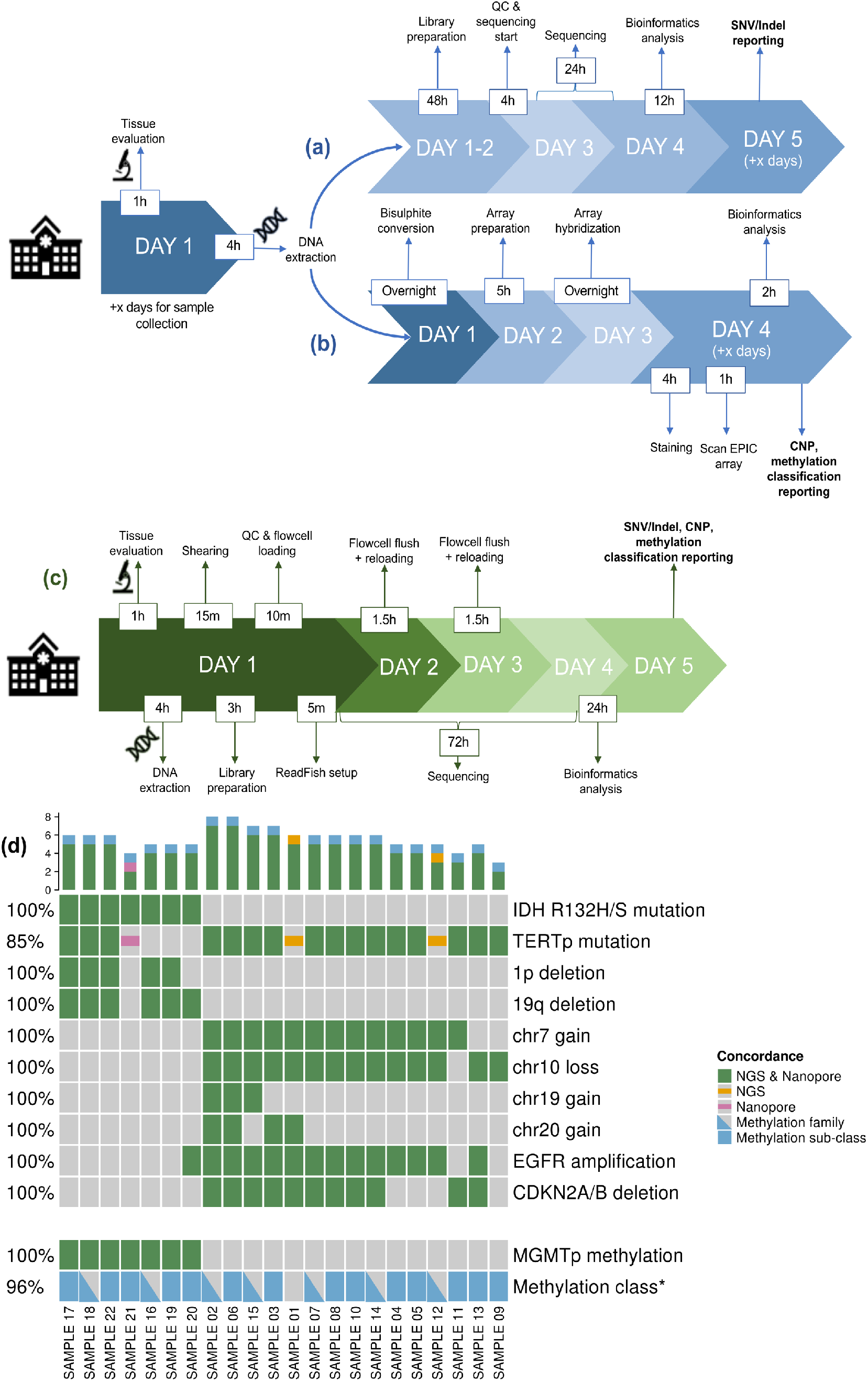
RAPID-CNS^2^ timeline and concordance. Timeline for (a) NGS panel sequencing and analysis pipeline, and (b) EPIC array analysis pipeline for neuropathology diagnostics (x denotes number of days required to pool sufficient samples). (c) Timeline for RAPID-CNS^2^ sequencing and analysis pipeline for a single sample. (d) Concordance of clinically relevant alterations & classification. Coloured blocks indicate presence of alteration, concordance for detected alterations is denoted in the legend. Triangular denotations for methylation class indicate samples where methylation families were concordant and blocks indicate concordance for sub-classes as well. Percentages on the left indicate concordance for the alteration over all samples.

Including CpG sites relevant for methylation-based classification in the ReadFish targets also allowed for methylation class prediction. The ability of nanopore sequencing to reliably provide a methylation classification using low-pass whole genome sequencing has previously been demonstrated by nanoDx [5]. Methylation families predicted by RAPID-CNS^2^ (the level most relevant for treatment decisions) matched their corresponding EPIC array-based classification in 21/22 cases, while precise methylation sub-classes were concordant in 14 cases. *MGMT* promoter status was also congruent with its corresponding EPIC array analysis for all cases [2]. Nanopore identified the *MGMT* promoter status as unmethylated in one sample in line with the EPIC array, which was assigned as methylated by pyrosequencing.

Targeted regions for RAPID-CNS^2^ can be easily altered by editing a BED file, in principle allowing lower sequencing times than in this study. With no additional library preparation steps required, it is possible to modify targeted regions for each individual sample as required. The MinION is a portable, handheld device which makes it a rational option for smaller neuropathology labs or in lower-infrastructure locations. While we used a GPU to run ReadFish, it can also be run using a sufficiently powerful CPU. Collectively, the RAPID-CNS^2^ approach can be set-up at low capital expense, is cost-efficient even in a low throughput setting, and provides a swift and highly flexible alternative to conventional NGS methods for SNV/InDel analysis, methylation classification and detection of copy number alterations.

## Data Availability

data will be made available upon reasonable request

## Acknowledgment

This study was supported by the Deutsche Forschungsgemeinschaft (DFG) via Comprehensive Research Center (SFB) 1389 Unite Glioblastoma.

## Supplementary methods

### Nanopore library prep optimized for adaptive sampling

Sections of 40×10 µm were prepared from cryoconserved tumor tissues with established molecular markers (IRB approval 2018-614N-MA, 005/2003) with tumor cell content (based on a H&E stain) > 60%. DNA was then extracted using the Promega Maxwell RSC Blood DNA Kit (catalogue # AS1400, Promega) on a Maxwell RSC 48 instrument (AS8500, Promega) per manufacturer’s instructions. DNA concentrations were measured on a microplate reader (FLUOStar Omega, BMG Labtech) using the Invitrogen Qubit DNA BR Assay Kit (Q32851, Thermo Fisher Scientific). Next, the DNA was sheared to approximately 9 to 11 kb in a total volume of 50 μl using g-TUBEs (Covaris) at 7200 rpm for 120 sec. The fragment length was assessed on an Agilent 2100 Bioanalyzer (catalogue # G2939A, Agilent Technologies) with the Agilent DNA 12000 Kit (catalogue # 5067- 1508, Agilent Technologies). Sequencing libraries were prepared with the SQK-LSK109 Ligation Sequencing Kit with the following modifications: 48 μl of the sheared DNA (2-2.5 μg) were taken into the end-prep reaction, leaving out the control DNA. The end-prep reaction was changed to an incubation for 30min at 20°C followed by 30min at 65°C followed by a cool down to 4°C in a thermal cycler. The clean-up was performed using AMPure XP beads and 80% ethanol, elution time was changed to 5min. Adapter ligation was extended to an incubation for 60min at room temperature. The ligation mix was then incubated with AMPure XP beads at 0.4x for 10min, clean- up was performed using the Long Fragment Buffer (LFB) and the final library was eluted in a total volume of 31 μl. Library concentrations were measured using the Invitrogen Qubit DNA HS Assay Kit (Q32851, Thermo Fisher Scientific) on a benchtop Quantus fluorometer (Promega). The libraries were loaded (500-600 ng) onto FLO- MIN106 R9.4.1 flow cells with a minimum of 1100 pores available according to the FC Check prior to loading. The flow cells were flushed after around 24 hours for a total of two times per sample with the Flow Cell Wash Kit (EXP-WSH003) per manufacturer’s instructions. All sequencing was carried out on a MinION 1B (Oxford Nanopore Technologies).

### ReadFish

Targeted nanopore sequencing was performed in real-time using a custom panel with ReadFish on an 8 GB NVIDIA RTX 2080 Ti powered consumer notebook [9]. The targets included regions from the neuropathology panel and CpG sites instrumental in classification by the random forest methylation classifier (available on GitHub). A 25kbp flank was added to the sites to ensure optimal targeting by ReadFish. Guppy 4.2.2’s fast basecalling (config dna_r9.4.1_450bps_fast) mode was used to run ReadFish.

### Bioinformatics analysis

FAST5 files were basecalled using Guppy 4.4.1’s high accuracy configuration. QC and coverage analyses were performed by pycoQC and deepTools respectively. Adapter trimming by Porechop was followed by minimap2 v2.18 alignment to the hg19 genome, samtools sorting and indexing. SNVs were called using longshot v0.4.1 and PEPPER-Margin-DeepVariant r0.4. TERT promoter mutations were detected by mpileup and bcftools. Variant annotation was performed by ANNOVAR. Filtering for clinical relevance was based on the 1000 Genomes (Aug 2015) frequencies and COSMIC 68 database. Copy number plots (100kb bin size) and gene-level copy number files (1kb, 10kb and 100kb bin sizes) were generated using CNVpytor and a custom script. Megalodon v2.3.1 was used to obtain methylation values.

### Methylation classification

To classify nanopore sequencing derived DNA-methylation profiles of central nervous system tumors, a random forest classifier was trained on publicly available 450k methylation array reference data set of the MNP classifier version 11 (GSE90496). This data set was preprocessed as described in [4].

For a batch of 22 nanopore sequencing samples, intersection of CpG probes measured for all samples were selected to train the classifier. The methylation array data set was reduced to these 3,285 probes.

Often nanopore sequencing measures CpG probes with low coverage, which leads to discrete distributed methylation values, i.e. (0, 0.5, 1) for coverage 3. As finer methylation differences can often not be detected with nanopore sequencing for all CpG probes, we trained the RF classifier on dichotomized methylation values. This followed the assumption that splitting rules learned on binary data are more robust and can be applied to methylation signals from nanopore sequencing data.

After dichotomizing the reduced reference methylation data set, a RF was trained with 1000 trees and the resulting permutation based variable importance measure was applied to select the 1000 CpGs with highest variable importance to train a final RF with again 1000 trees. The out-of-the-bag accuracy of this classifier was 96%.

## Supplementary results

### RAPID-CNS^2^ analysis pipeline

The bioinformatics pipeline requires raw FAST5 files as input. Complete instructions for setting up the analysis are available on GitHub. Post set-up, RAPID-CNS^2^ runs the entire analysis with a single command. It can be run on an LSF cluster or a GPU workstation. Basecalling followed by SNV and CNV detection completes within 10 hours while methylation calling and classification requires an additional 12 hours.

### SNV detection

ANNOVAR annotated tables for all Nanopore sequenced samples and their corresponding panel sequencing results are attached.

### CNV detection

Copy number plots obtained using the RAPID-CNS^2^ pipeline demonstrate higher resolution and clear visualization of the copy number levels as compared to NGS panel sequencing (Supplementary figure 1a (left and centre)). Calculating depth of mapped reads, copy number variations detected are comparable to EPIC array results (Supplementary figure 1a (left and right)). Normalised read depths are indicated on the Y-axis with “2” indicating mean autosomal level. Additionally, genes covered by the copy number variations and their zygosity are annotated and output as excel files (Supplementary files).

### MGMT promoter methylation

Two probes used by the MGMT-STP27 approach were not reliably covered in all analysed samples [2]. Methylation frequencies over all CpG sites covering the *MGMT* promoter region were therefore averaged as an alternative measure. Using pyrosequencing as gold standard, methylated and unmethylated samples were found to have a significant difference in their average methylation (Wilcoxon rank sum test p-value= 2.719e-06). As shown in Suppl. Figure 1b, a threshold of 10% was assigned for *MGMT* promoter methylation status.

**Supplementary Figure 1:**
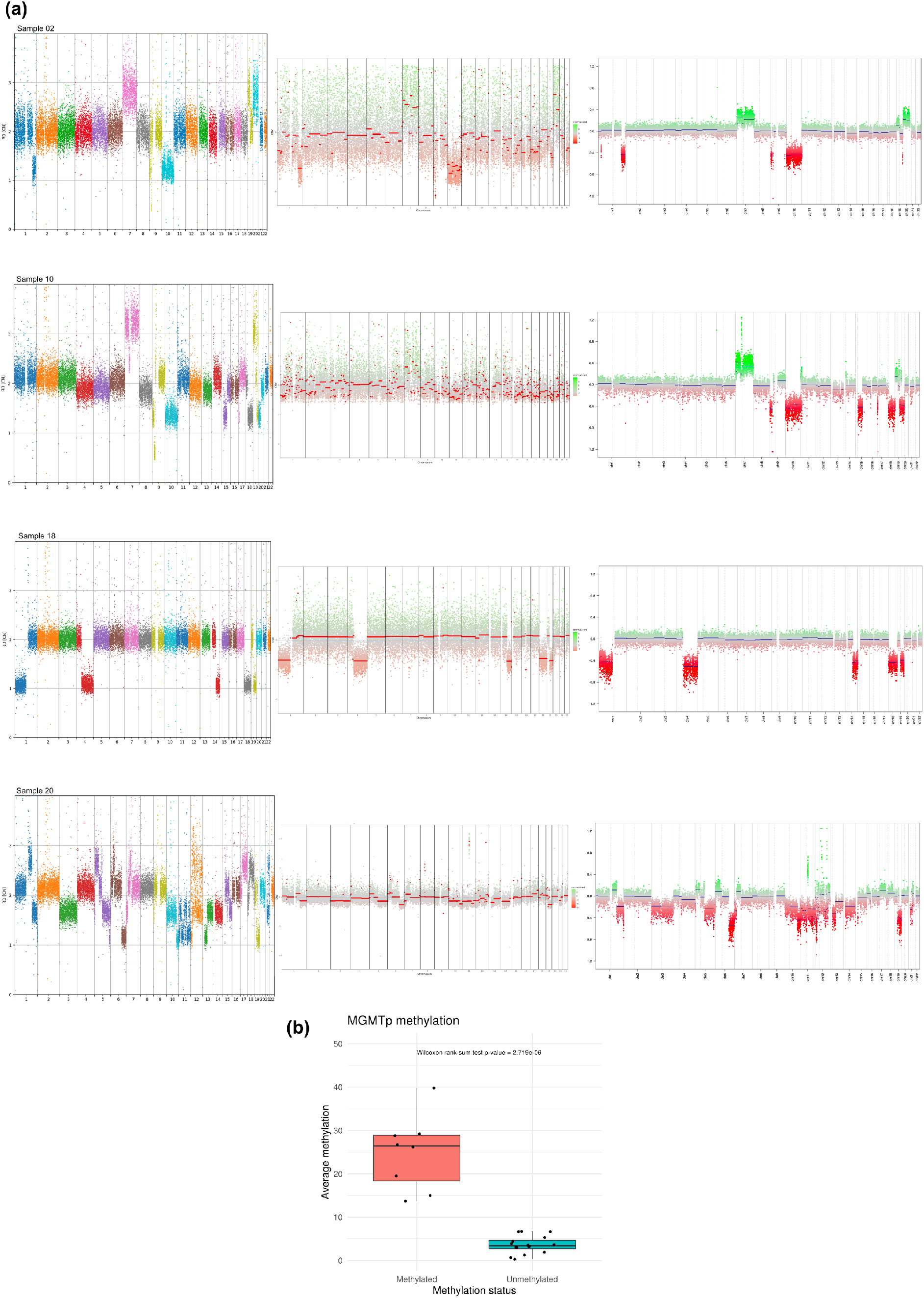
(a) CNV plots obtained using RAPID-CNS^2^ (left), panel-sequencing (centre) and EPIC array analysis (right). (b) MGMT promoter methylation values averaged over the MGMT promoter region.

